# Neural Network Predicts Need for Red Blood Cell Transfusion for Patients with Acute Gastrointestinal Bleeding Admitted to the Intensive Care Unit

**DOI:** 10.1101/2020.05.19.20096743

**Authors:** Dennis Shung, Egbert Castro, Jessie Huang, J. Kenneth Tay, Michael Simonov, Loren Laine, Smita Krishnaswamy

## Abstract

**Background:** Acute gastrointestinal bleeding is the most common gastrointestinal cause for hospitalization. For high risk patients requiring intensive care unit stay, predicting transfusion needs during the first 24 hours using dynamic risk assessment may improve resuscitation.

**Aims:** Provide dynamic risk prediction for red blood cell transfusion in admitted patients with severe acute gastrointestinal bleeding.

**Methods:** A patient cohort admitted for acute gastrointestinal bleeding (N = 2,524) was identified from the Medical Information Mart for Intensive Care III (MIMIC-III) critical care database, separated into training (N = 2,032) and validation (N = 492) sets. 74 demographic, clinical, and laboratory test features were consolidated into 4-hour time intervals over the first 24 hours from admission. The outcome measure was the transfusion of red blood cells during each 4-hour time interval. A long short-term memory (LSTM) model, a type of Recurrent Neural Network (RNN), was compared to the Glasgow-Blatchford Score (GBS).

**Results:** The LSTM model performed better than GBS overall (AUROC 0.81 vs 0.63*;P*<0.001) and at each 4-hour interval (*P*<0.01). At high sensitivity and high specificity cutoffs, the LSTM model outperformed GBS (*P*<0.001). The LSTM model performed better in patients directly admitted from the ED to ICU (0.82 vs 0.63;*P*<0.001), upper GIB (0.84 vs 0.68;*P*<0.001), lower GIB (0.77 vs 0.58;*P*<0.001), and unspecified GIB (0.85 vs 0.64;*P*<0.001).

**Conclusions:** A LSTM model can be used to predict the need for transfusion of packed red blood cells over the first 24 hours from admission to help personalize the care of high-risk patients with acute gastrointestinal bleeding.

**Data Access:** All clinical data from MIMIC-III was approved under the oversight of the Institutional Review Boards of Beth Israel Deaconess Medical Center (Boston, MA) and the Massachusetts Institute of Technology (Cambridge, MA). Requirement for individual patient consent was waived because the project did not impact clinical care and all protected health information was deidentified. The data was available on PhysioNet were derived from protected health information that has been de-identified and not subject to HIPAA Privacy Rule restrictions. All use of the data was performed with credentialed access under the oversight of the data use agreement through PhysioNet and the Massachusetts Institute of Technology.

## Introduction

Acute gastrointestinal bleeding accounts for over 2.2 million hospital days and 19.2 billion dollars of medical charges annually in the United States and frequently requires red-blood cell transfusion.^1^ The management of severe acute gastrointestinal bleeding begins with resuscitation using intravenous fluids and transfusion of packed red blood cells, which are given to 43% of patients hospitalized with upper gastrointestinal bleeding in the United Kingdom and 21% of patients hospitalized with lower gastrointestinal bleeding in the United States.^2,3^

Transfusion needs may change during the hospital stay, but a tool to dynamically predict transfusion needs over time does not yet exist in clinical care. Patients with severe acute gastrointestinal bleeding who require care in the intensive care setting generally have higher transfusion needs and may benefit most from a predictive tool to guide resuscitation efforts. Current guidelines are based on a restrictive transfusion strategy using a hemoglobin threshold for transfusion of 7g per deciliter, which has been associated with a significant 45% relative risk reduction in 45-day mortality as compared to a threshold of 9g per deciliter in patients with upper gastrointestinal bleeding.^4^

Dynamic risk prediction, where predictions are generated in real time every hour based on clinical and laboratory values, may help guide resuscitation strategies and help in timing endoscopic intervention, particularly in severely ill patients who require intensive care. Existing clinical risk scores used to screen for risk of needing transfusion of packed red blood cells, such as the Glasgow-Blatchford Score, are static models that only use clinical information at the time of admission (e.g. initial systolic blood pressure).^5^ Machine learning approaches to model risk for gastrointestinal bleeding have shown promise in outperforming existing clinical risk scores, but are also static models.^6,7^ Electronic health records (EHRs) can capture clinical data in real time, and have been used to create automated tools to model adverse events, such as sepsis, post-operative complications, and acute kidney injury.^8-11^ Neural networks, computing systems inspired by the human brain, consisting of collections of layered neurons that process and transmit information to the next layer of neurons, are a transformative machine learning model that have the potential to dynamically predict the need for blood transfusion for patients over time.^12^ Recurrent neural networks have been demonstrated to be better than state-of-the-art risk models for continuous prediction of acute kidney injury up to 48 hours, the onset of septic shock 28 hours before onset, and all-cause inpatient mortality.^13-15^ We propose the use of a Long-Short-Term Memory (LSTM) Network, an advanced recurrent neural network that learns what to store in its memory, to process data from electronic health records with an internal memory that stores relevant information over time and can generate a probability of transfusion within the 4 hour intervals for patients with severe acute gastrointestinal bleeding. Figure 1 shows the use of our model in an example patient from the MIMIC test set with generated risk predictions throughout the first 24 hours from admission. (Figure 1)

**Figure 1:**
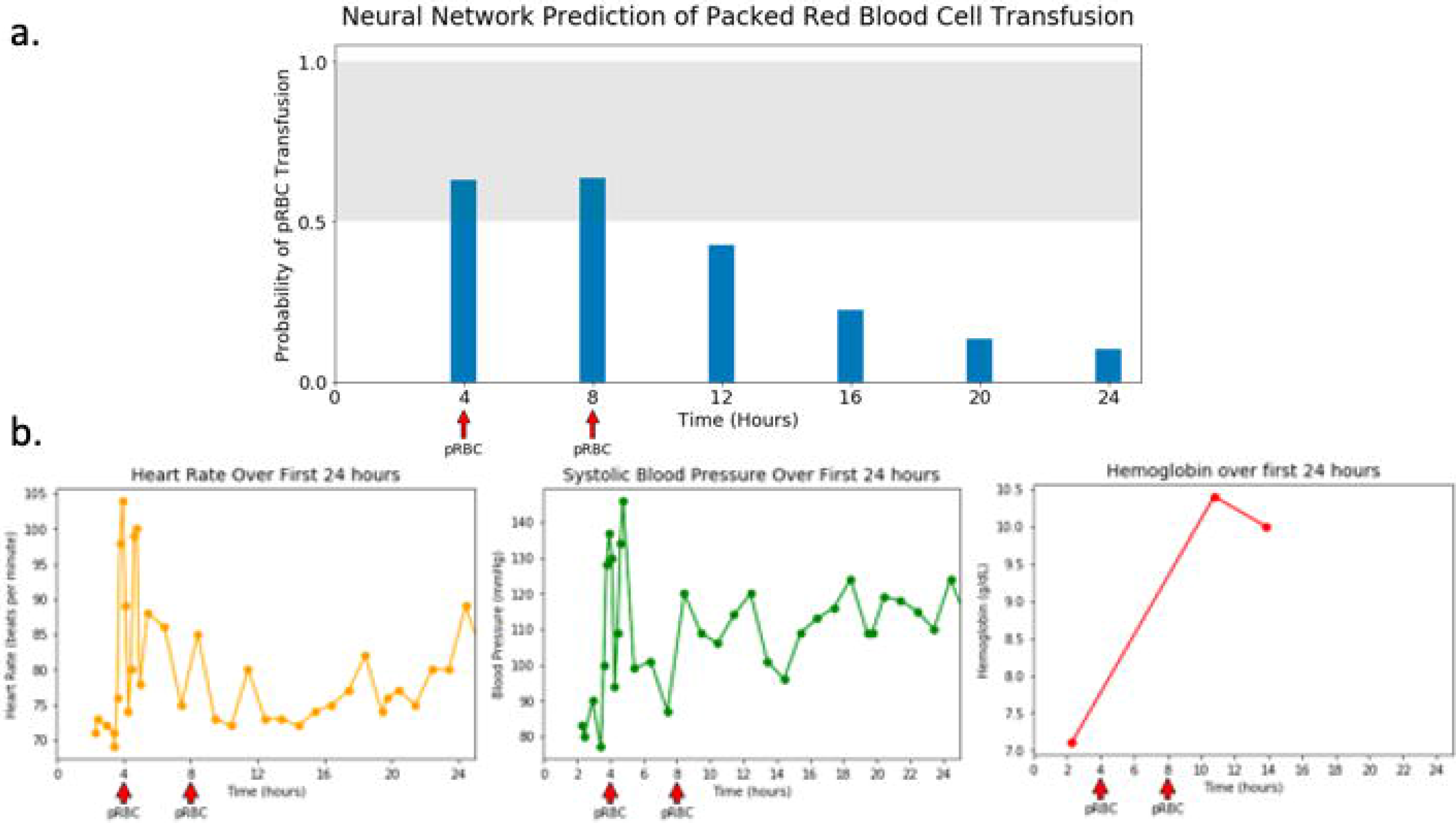
Example of neural network prediction for the first 24 hours of a 62 year old man with Hepatitis C cirrhosis presenting with 2 days of intermittent coffee ground emesis and lethargy. Glasgow Blatchford Score = 14 a) Continuous risk prediction of the neural network through the first 24 hours with the threshold set above 0.5 for detecting need for transfusion. The arrows indicate need for transfusion during that time period. b) Measurements of Heart Rate, Systolic Blood Pressure, and Hemoglobin occurring during the first 24 hours.

## Methods

### Data Source

A patient cohort presenting with acute gastrointestinal bleeding was identified from the Medical Information Mart for Intensive Care III (MIMIC-III) critical care database.^16,17^ The database contains data for over 40,000 patients in the Beth Israel Deaconess Medical Center from 2001 to 2012 requiring an ICU stay.

Patients were included if they had an admission diagnosis containing the terms “gastro”, “bleed”, “melena”, “hematochezia”. The diagnoses were collated and then manually reviewed. Patients were excluded if vital signs were only available greater than 24 hours from time of admission. The data included information that was updated over time during the course of hospitalization, including laboratory results and vital signs. For laboratory values, any negative entry or non-quantizable (e.g. >=, <) was converted to missing. Medications, current procedural terminology codes, and ICD9/10 codes from the visit were excluded from the analysis. The dataset had a total of 74 features: 5 clinical and demographic variables and 69 laboratory variables.

### Study Design

The final cohort included 2,524 hospital admissions and was randomly split into a training set with 2,032 hospital admissions and a validation set with 492 hospital admissions. (Table 1) We chose to compare the model to the Glasgow-Blatchford Score, which was designed to predict need for transfusion of packed red blood cells or intervention to control bleeding, rebleeding, or dying.^5^ A Glasgow-Blatchford Score was calculated through manual review of Admission Notes extracted from MIMIC-III for all patients in the validation set. The performance of the neural network model and the GBS were compared by applying the same GBS for each 4-hour time interval for the first 24 hours and comparing the neural network output at each 4-hour interval.

**Table 1:** Demographics and Baseline Data for the Training and Validation Set

|  | Training Set<br>N = 2,032 |  | Validation Set<br>N = 492 |  |  |
| --- | --- | --- | --- | --- | --- |
|  | N | Proportion | N | Proportion | p-value |
| **Demographic Information** |  |  |  |  |  |
| Male | 836 | 41% | 190 | 39% | 0.31 |
| Age |  |  |  |  |  |
| >89 | 144 | 7% | 42 | 9% | 0.29 |
| 75-89 | 629 | 31% | 168 | 34% | 0.24 |
| 50-75 | 935 | 46% | 200 | 41% | 0.14 |
| 25-50 | 316 | 16% | 71 | 14% | 0.43 |
| <25 | 8 | 0% | 4 | 1% | 0.27 |
| Ethnicity |  |  |  |  |  |
| White | 1429 | 70% | 380 | 77% | 0.08 |
| African American | 244 | 12% | 52 | 11% | 0.35 |
| Hispanic | 75 | 4% | 22 | 4% | 0.37 |
| Asian American | 74 | 4% | 15 | 3% | 0.42 |
| Other | 210 | 10% | 23 | 5% | 0.05 |
| **Clinical Features** |  |  |  |  |  |
| Upper Gastrointestinal Bleeding | 679 | 33% | 203 | 41% | 0.07 |
| Lower Gastrointestinal Bleeding | 428 | 21% | 162 | 33% | 0.02 |
| Unspecified Location | 925 | 46% | 127 | 26% | <0.01 |
| **Outcomes** |  |  |  |  |  |
| Packed Red Blood Cells | 1542 | 76% | 381 | 77% | 0.39 |
| In-Hospital Mortality | 156 | 8% | 32 | 6.5% | 0.35 |
|  | Mean | Std Dev | Mean | Std Dev | p-value |
| **Vital Signs** |  |  |  |  |  |
| Heart Rate (beats per minute) | 88.9 | 18 | 88.1 | 16.6 | 0.35 |
| Systolic Blood Pressure | 126.9 | 22.9 | 127.1 | 22.2 | 0.86 |
| Diastolic Blood Pressure | 64.2 | 16.9 | 65.7 | 16.6 | 0.07 |
| **Laboratory Tests** |  |  |  |  |  |
| Alanine Aminotransferase (ALT) | 41 | 86.9 | 39.8 | 152 | 0.87 |
| Albumin | 3.1 | 0.63 | 3.1 | 0.65 | 1.00 |
| Alkaline Phosphatase | 117.2 | 152.8 | 118.4 | 130.5 | 0.86 |
| Anion Gap | 15 | 4.3 | 14.8 | 4 | 0.33 |
| Aspartate Aminotransferase (AST) | 75.8 | 159 | 75 | 446 | 0.97 |
| Bicarbonate | 23.8 | 4.5 | 24 | 4.4 | 0.37 |
| Bilirubin, Total | 1.52 | 3 | 1.47 | 2.5 | 0.70 |
| Calcium, Total | 8.1 | 0.88 | 8.2 | 0.8 | 0.01 |
| Chloride | 104.4 | 6.1 | 104.4 | 6.5 | 1.00 |
| Creatinine | 1.43 | 1.2 | 1.5 | 1.4 | 0.31 |
| Glucose | 147 | 77.5 | 143.8 | 60.6 | 0.32 |
| Magnesium | 1.9 | 0.71 | 1.9 | 0.4 | 1.00 |
| Phosphate | 3.5 | 1.2 | 3.5 | 1.2 | 1.00 |
| Potassium | 4.3 | 0.76 | 4.4 | 0.8 | 0.01 |
| Sodium | 138.8 | 4.6 | 138.7 | 4.8 | 0.68 |
| Urea Nitrogen | 37.5 | 28 | 39.5 | 30.4 | 0.18 |
| Basophils | 0.39 | 0.98 | 0.34 | 0.34 | 0.06 |
| Eosinophils | 1.4 | 1.9 | 1.4 | 1.7 | 1.00 |
| Hematocrit | 28 | 9.5 | 27.7 | 6.5 | 0.41 |
| Hemoglobin | 9.5 | 2.5 | 9.3 | 2.4 | 0.10 |
| International Normalized Ratio (INR) | 1.8 | 2.5 | 1.8 | 1.7 | 1.00 |
| Lymphocytes (%) | 17.1 | 10.2 | 16.6 | 10.1 | 0.33 |
| Mean Corpuscular Hemoglobin (MCH) | 30.2 | 3 | 29.9 | 2.9 | 0.04 |
| Mean Corpuscular Hemoglobin Concentration (MCHC) | 33.5 | 1.7 | 33.3 | 1.8 | 0.03 |
| Mean Corpuscular Volume (MCV) | 90 | 7.6 | 89.7 | 7.3 | 0.42 |
| Monocytes (%) | 4.6 | 2.6 | 4.6 | 2.3 | 1.00 |
| Neutrophils (%) | 75.1 | 12.3 | 76.5 | 11.7 | 0.02 |
| Platelet Count (x1000) | 231.8 | 139.3 | 238.9 | 127.4 | 0.28 |
| Prothrombin Time (PT) | 17.1 | 11.4 | 19.1 | 15.7 | 0.01 |
| Partial Thromboplastin Time (PTT) | 31.3 | 13.7 | 31.3 | 14.2 | 1.00 |
| Red Blood Cell Distribution Width (RDW) | 16.1 | 2.5 | 15.9 | 2.2 | 0.08 |

### Comparison of High Sensitivity and High Specificity Thresholds

Pre-defined cutoff of >90% was defined for sensitivity and specificity of the LSTM neural network model and the Glasgow Blatchford Score. A high sensitivity threshold means that the algorithm would not miss time periods when patients need packed red blood cell transfusions. A high specificity threshold means that the algorithm would only indicate need for packed red blood cell transfusion during a time period where the patient actually needs one, minimizing false alerts. The specific high sensitivity and high specificity threshold for the Glasgow Blatchford Score was matched to the corresponding cutoff for the LSTM Neural Network.

### Sensitivity Analysis

Patients who are admitted directly from the emergency department to the intensive care unit may be different from patients who are transferred to the ICU from an inpatient ward. We applied our model to a validation set with only patients who were admitted to the ICU directly from the emergency department, as defined by arrival to ICU less than 1 hour from leaving the emergency department. We also perform this sensitivity analysis on subsets of patients presenting with upper GIB, lower GIB, and GIB of unspecified source.

### Input Variables

A total of 74 input variables were used and included age, gender, vital signs (systolic blood pressure, diastolic blood pressure, heart rate), and 69 unique laboratory values. (Table 2) The vital signs and laboratory values were extracted and then consolidated into 4-hour time intervals over the first 24 hours from admission.

**Table 2:**
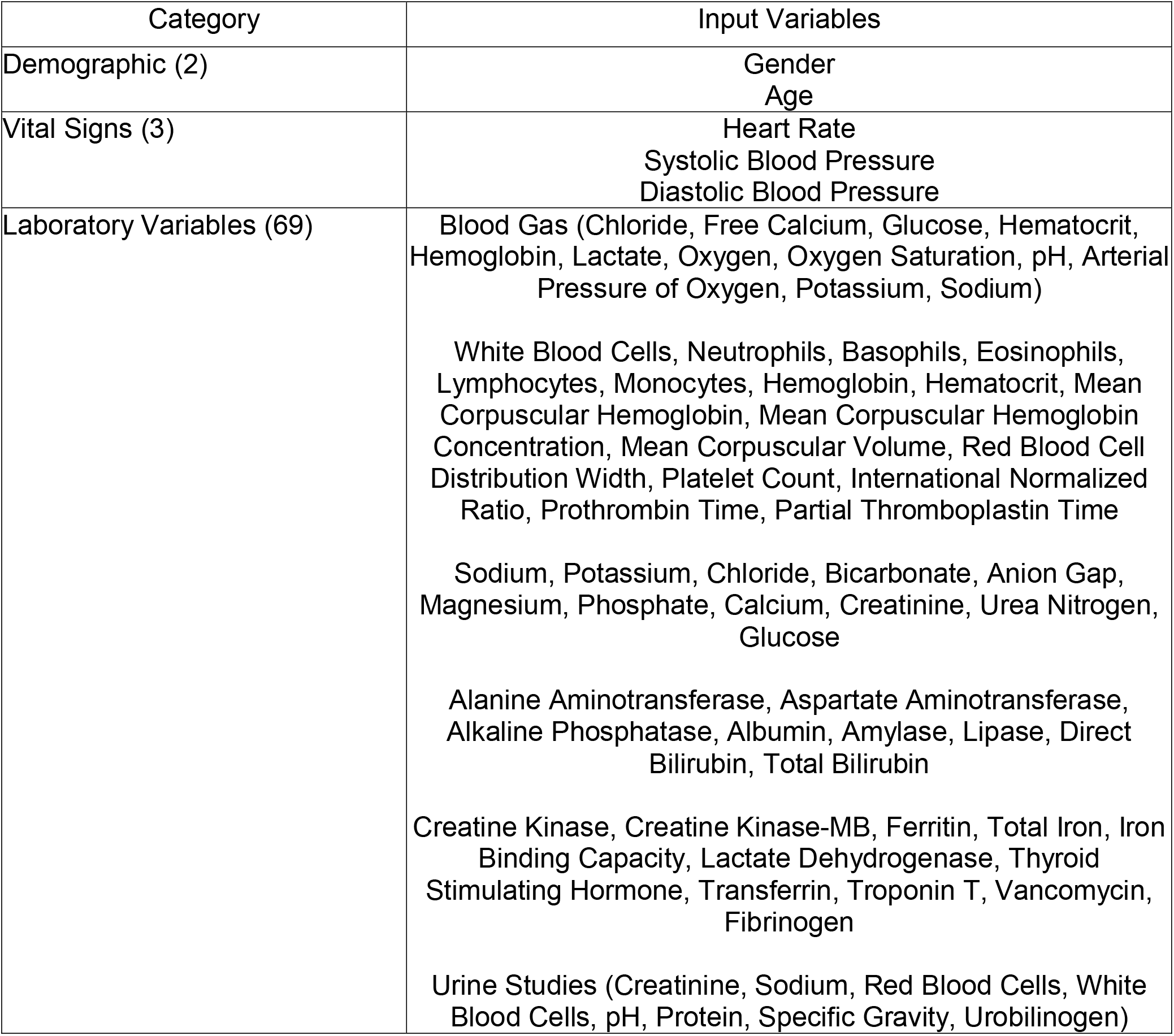
Input Variables (N = 74)

| Category | Input Variables |
| --- | --- |
| Demographic (2) | Gender<br>Age |
| Vital Signs (3) | Heart Rate<br>Systolic Blood Pressure<br>Diastolic Blood Pressure |
| Laboratory Variables (69) | Blood Gas (Chloride, Free Calcium, Glucose, Hematocrit, Hemoglobin, Lactate, Oxygen, Oxygen Saturation, pH, Arterial Pressure of Oxygen, Potassium, Sodium)<br>White Blood Cells, Neutrophils, Basophils, Eosinophils, Lymphocytes, Monocytes, Hemoglobin, Hematocrit, Mean Corpuscular Hemoglobin, Mean Corpuscular Hemoglobin Concentration, Mean Corpuscular Volume, Red Blood Cell Distribution Width, Platelet Count, International Normalized Ratio, Prothrombin Time, Partial Thromboplastin Time<br>Sodium, Potassium, Chloride, Bicarbonate, Anion Gap, Magnesium, Phosphate, Calcium, Creatinine, Urea Nitrogen, Glucose<br>Alanine Aminotransferase, Aspartate Aminotransferase, Alkaline Phosphatase, Albumin, Amylase, Lipase, Direct Bilirubin, Total Bilirubin<br>Creatine Kinase, Creatine Kinase-MB, Ferritin, Total Iron, Iron Binding Capacity, Lactate Dehydrogenase, Thyroid Stimulating Hormone, Transferrin, Troponin T, Vancomycin, Fibrinogen<br>Urine Studies (Creatinine, Sodium, Red Blood Cells, White Blood Cells, pH, Protein, Specific Gravity, Urobilinogen) |

### Outcome Variable

The predicted outcome measure was the transfusion of packed red blood cells, calculated as binary 0 (no transfusion) or 1 (transfusion given). At the beginning of each 4-hour time interval, the model makes a prediction on whether a transfusion will be needed by the end of the 4-hour interval.

### Data Pre-Processing

Each patient was represented by a sequence of events with each 4-hour period containing information recorded in the vitals and laboratory values. Information for each patient was encoded into 4-hour time intervals up to the first 24 hours. After excluding lab values with greater than 90% missingness, remaining lab values with greater than 50% missingness in the dataset were converted to missing indicator variables, with 1 as present and 0 as missing. To harmonize the input variables across patients, the first timepoint for each patient was fixed at the first recording of heart rate, systolic blood pressure, and diastolic blood pressure. Consolidation of vital signs and laboratory values in each 4-hour interval was performed by taking the mean of each value. All continuous values were normalized and centered. Age was maintained as a continuous variable, with patients greater than 89 years old coded as 89 years old.

### Missing Values

To examine the role of the data imputation method used, we compared 4 different imputation strategies. The first was imputation of the mean value of for any missing value. The second was a carryforward approach, or using the previously recorded value if a value was present at a previous time point but no subsequent value was measured. This assumes that the laboratory value is constant until the next time point in clinical decision-making.^18^ The third was mean imputation with a new variable that served as a missingness indicator for every variable. The fourth was carryforward with a missingness indicator for every variable.

### Model background

Recurrent neural networks allow for processing of sequential information by storing information as internal states over multiple time points. Long short-term memory (LSTM) networks are a type of RNN that can be useful for clinical measurements because they carefully tune the information passed between subsequent time-iterations of the model. Given a series of HER data, ***x***^(**0**)^, ***x***^(**1**)^,…, ***x***^(***T***-**1**)^, where ***x***^(***t***)^ represents the input variables for the (*t* + 1)th 4-hour interval, at the beginning of each 4-hour interval our goal is to predict whether transfusion is needed in the next 4 hours. The output is a sequence of probability predictions 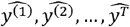, where 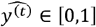 is the prediction for whether transfusion is needed in the t^th^ 4-hour interval. The LSTM model consists of 2 layers of 128 LSTM cells each, followed with a linear layer that maps from hidden state space to the prediction space. We obtain the log-probabilities by adding a LogSoftmax later in the last layer of the network. Thus the output of the neural network is a sequence 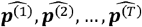, where 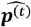 is the log-probability of 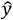 being either of the target classes, and our decision rule is to administer transfusion if 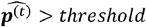, where the threshold is determined by desired sensitivity or specificity. We use the negative log likelihood for the output at each time of interest as the loss function. The model is trained for up to 100 epochs with hyperparameters corresponding to the lowest validation loss recorded and used to obtain testing accuracy.

### Statistical Analysis

Two-tailed t tests and chi-squared test were used to compare baseline characteristics between the training and validation sets. We assessed model performance using the area under the curve (AUROC) and compared it to the performance of GBS using the nonparametric DeLong test.^19^ McNemar’s test was used to compare specificity and sensitivity at specified cutoffs between the two risk assessment models.

## Results

Demographics were similar between training and validation sets with the median age 69 for both, proportion of men (41% in training, 39% on validation), and predominantly white (70% in training, 77% in validation). Both sets had similar percentage of patients with upper gastrointestinal bleeding (training 33% vs validation 41%). The training set had more patients with gastrointestinal bleeding and unspecified source (46% vs 26% *P*<0.01), while the validation set had more patients with lower gastrointestinal bleeding (33% vs 21% *P*=0.02). Vital signs and laboratory values were similar in the training and validation sets. (Table 1)

The performance of the LSTM model on the four different imputation strategies were similar and all significantly better than GBS. (Table 5) The results we subsequently present are for the strategy with the highest AUROC (mean imputation and missing indicators). For the main analysis of all patients with acute gastrointestinal bleeding who were transferred to the ICU, the LSTM performed significantly better than GBS in predicting packed red blood cell transfusion across the entire 24 hour period (AUROC 0.81 CI 0.80-0.83 vs 0.63 CI 0.61-0.65;*P*<0.001). The model also consistently outperformed GBS in each 4-hour interval. (Table 3, Figure 2) At the cutoff for high specificity (>90%), the LSTM performed better than GBS (LSTM sensitivity 42% versus GBS sensitivity 12%;*P*<0.001). At the cutoff for high sensitivity (>90%), the LSTM performed better than GBS (LSTM sensitivity 37% versus GBS sensitivity 15%;*P*<0.001). (Table 6)

**Table 3:** Performance of the Long-Short Term Memory (LSTM) Model and the Glasgow Blatchford Score in Predicting Transfusion of Packed Red Blood Cells by Comparison of Area Under the Receiver Operating Curve (AUROC) for all patients (N = 492) overall and by each 4 hour interval.

|  | Long-Short Term Memory<br>Network Model<br>Validation<br>AUROC<br>95% Confidence Interval | Glasgow-Blatchford<br>Score<br>Validation<br>AUROC<br>95% Confidence<br>Interval | p-value |
| --- | --- | --- | --- |
| **Overall** | **0.81**<br>**(0.80-0.83)** | **0.63**<br>**(0.61-0.65)** | **<0.001** |
| Hour 4 | 0.79<br>(0.75-0.83) | 0.70<br>(0.65-0.74) | <0.001 |
| Hour 8 | 0.76<br>(0.72-0.80) | 0.64<br>(0.59-0.69) | <0.001 |
| Hour 12 | 0.72<br>(0.67-0.77) | 0.62<br>(0.56-0.67) | <0.001 |
| Hour 16 | 0.73<br>(0.68-0.79) | 0.65<br>(0.59-0.71) | 0.005 |
| Hour 20 | 0.78<br>(0.72-0.84) | 0.62<br>(0.55-0.69) | <0.001 |
| Hour 24 | 0.78<br>(0.72-0.84) | 0.60<br>(0.52-0.69) | <0.001 |

**Table 4:** Sensitivity Analyses of Overall Performance of the Long-Short Term Memory (LSTM) Model and the Glasgow Blatchford Score in Predicting Transfusion of Packed Red Blood Cells by Comparison of Area Under the Receiver Operating Curve (AUROC)

|  | Long-Short Term Memory<br>Network Model<br>Validation<br>AUROC<br>95% Confidence Interval | Glasgow-Blatchford<br>Score<br>Validation<br>AUROC<br>95% Confidence<br>Interval | p-value |
| --- | --- | --- | --- |
| Patients from<br>ED to ICU<br>(N = 412) | 0.82<br>(0.80-0.83) | 0.63<br>(0.61-0.66) | <0.001 |
| Upper GIB<br>(N = 203) | 0.84<br>(0.81-0.86) | 0.68<br>(0.64-0.71) | <0.001 |
| Lower GIB<br>(N = 162) | 0.77<br>(0.74-0.80) | 0.58<br>(0.54-0.61) | <0.001 |
| GIB with<br>Unspecified<br>Source<br>(N = 127) | 0.85<br>(0.82-0.88) | 0.64<br>(0.59-0.68) | <0.001 |

**Table 5:**
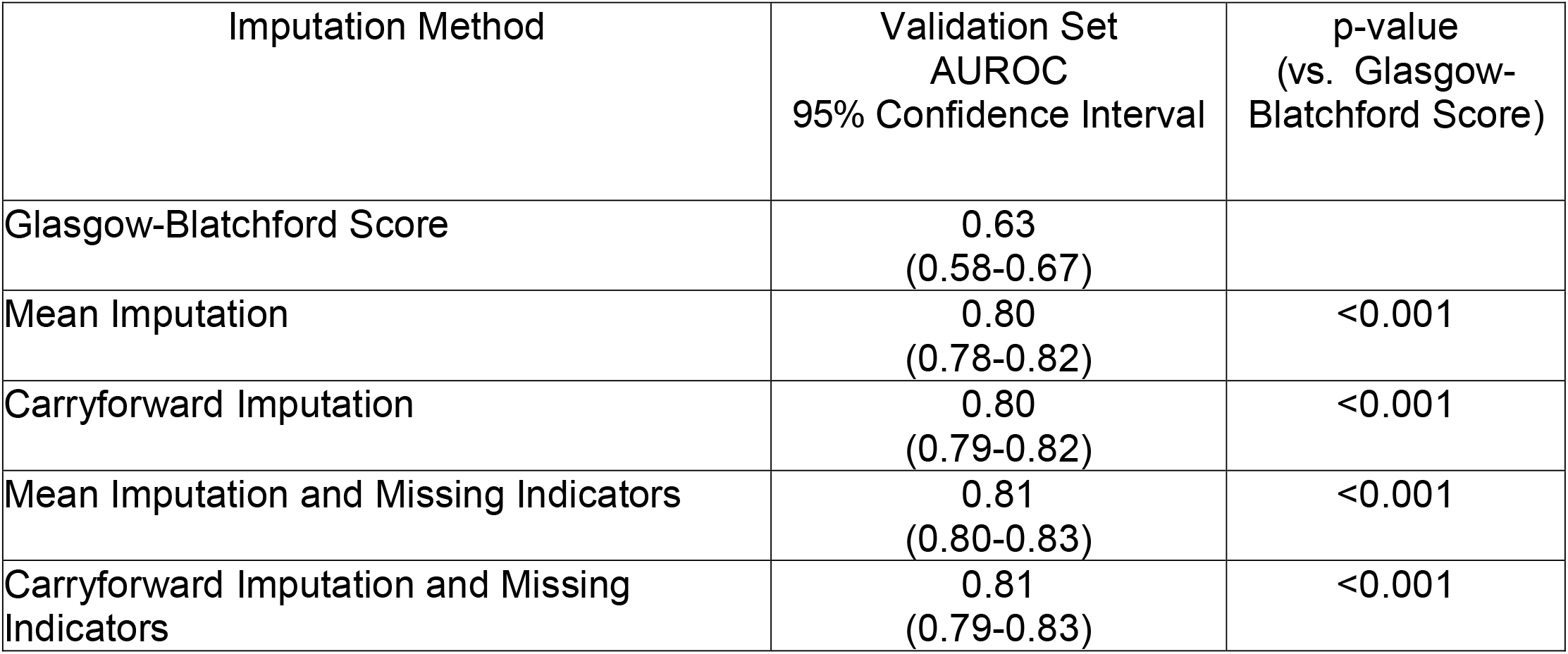
Comparison of the overall performance of Long-Short Term Memory network model with different imputation methods to address missingness in the first 24 hours after admission for all patients admitted to the Intensive Care Unit with Acute Gastrointestinal Bleeding. The Glasgow-Blatchford Score was able to be calculated with no missing values at presentation.

| Imputation Method | Validation Set<br>AUROC<br>95% Confidence Interval | p-value<br>(vs. Glasgow-Blatchford Score) |
| --- | --- | --- |
| Glasgow-Blatchford Score | 0.63<br>(0.58-0.67) |  |
| Mean Imputation | 0.80<br>(0.78-0.82) | <0.001 |
| Carryforward Imputation | 0.80<br>(0.79-0.82) | <0.001 |
| Mean Imputation and Missing Indicators | 0.81<br>(0.80-0.83) | <0.001 |
| Carryforward Imputation and Missing Indicators | 0.81<br>(0.79-0.83) | <0.001 |

**Table 6:** Performance of LSTM Neural Network model Compared to the Glasgow Blatchford Score at matched cutoffs of 91% sensitivity and 93% for the internal validation dataset

| **Risk Stratification Tool** | **Sensitivity** | **Specificity** |
| --- | --- | --- |
| *High Specificity Cutoff* |  |  |
| Glasgow Blatchford Score = 14 | 12% | 93% |
| LSTM Neural Network | 42% | 93% |
| *High Sensitivity Cutoff* |  |  |
| Glasgow Blatchford Score = 5 | 91% | 22% |
| LSTM Neural Network | 91% | 46% |

**Figure 2:**
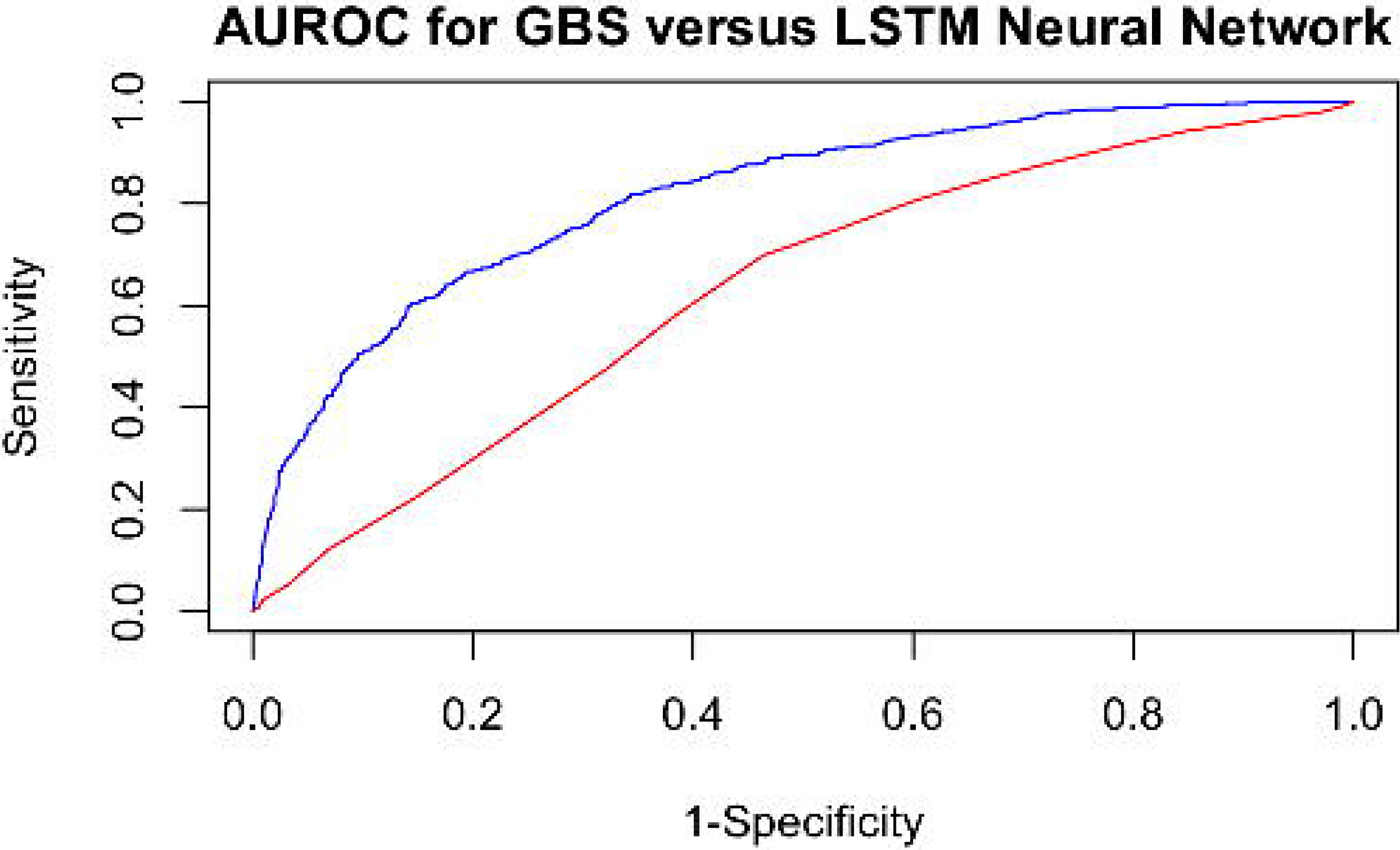
Comparison of the overall Area Under the Receiver Operating Curve (AUROC) as a measure of performance of the Long-Short Term Memory (LSTM) Neural Network model and Glasgow-Blatchford Score (GBS).

For the sensitivity analysis of only patients admitted directly from the ED to the ICU, the LSTM network was also significantly better than GBS (AUROC 0.82 CI 0.80-0.83 vs. 0.63 CI 0.61-0.66, *P*<0.001). The LSTM network was also significantly better than GBS for upper GIB (0.84 CI 0.81-0.86 vs 0.68 CI 0.64-0.71;*P*<0.001), lower GIB (0.77 CI 0.74-0.80 vs 0.58 CI 0.54-0.61;*P*<0.001), and GIB of unspecified origin (0.85 CI 0.82-0.88 vs 0.64 CI 0.59-0.68;*P*<0.001). (Table 4) There was no clear difference in performance as measured by AUROC in patients who were directly admitted to the ICU from the ED.

## Discussion

Predicting the need for transfusion of packed red blood cells has direct relevance to guiding the management of patients with acute gastrointestinal bleeding. This is the first study to show that a LSTM network model is able to predict the need for packed red blood cell transfusion for patients with severe acute gastrointestinal bleeding. By anticipating needs for transfusion, this is a first step towards personalizing treatment and tailoring appropriate resuscitation to reduce clinical decompensation and death for patients with severe acute gastrointestinal bleeding. While endoscopic evaluation is important, adequate resuscitation is the cornerstone of management prior to endoscopy.^20-23^

Previous risk scores capture information from specific points in time at admission, and do not incorporate new clinical data over the course of hospitalization. Electronic health records contain longitudinal information on patients admitted to the hospital and reflect real-world practice, which can be used to develop risk prediction models.^24^ For patients who have severe disease requiring intensive care unit stay, mortality may be more due to end organ damage due to inadequate perfusion.^3,25,26^ Despite the significant computing requirements necessary to run neural networks, existing electronic health records are now deploying cloud computing infrastructure able to perform computationally intensive tasks. The emerging capabilities of cloud infrastructure in electronic health records, such as the Cognitive Computing platform for Epic Systems, make the deployment of neural networks for clinical care feasible.

We envision the future of care for all patients to be enhanced by customized machine learning decision support tools that will provide both initial risk stratification and ongoing risk assessment to provide treatment at the right time for the right patient. Using a dynamic risk assessment, resuscitation needs could be estimated early and optimized in preparation for endoscopic evaluation and intervention. This individualized decision-making potentially will minimize organ damage from inadequate resuscitation, which drives the risk for mortality in these patients.^25^ The LSTM model can be tuned for provider preference with either a high sensitivity threshold or a high specificity threshold. Alert fatigue is particularly relevant in the ICU, since clinically irrelevant alerts can have an impact on patient safety.^27^ In order to minimize alert fatigue, a high specificity threshold could be set for the algorithm. However, if providers do not want to miss any time periods when patients need packed red blood cell transfusions, a high sensitivity threshold can be set to minimize false negatives. Although the LSTM network model is much better than a currently used tool (GBS), it still falls short of optimal performance. More work will be needed to develop and validate neural network models.

Strengths of this study include modeling patients with severe illness requiring intensive care unit stay, which may benefit disproportionately from timely transfusion and resuscitation and the use of vital signs and laboratory tests that are standardized and can be easily mapped across electronic health record systems.

Limitations include the absence of prospective and independent validation in other datasets and proportion of missing data that required imputation methods, which may introduce bias to the data. Furthermore, our comparison to Glasgow-Blatchford Score may be suboptimal, since we studied patients with any overt gastrointestinal bleeding regardless of source, but the GBS was primarily developed for patients with upper gastrointestinal bleeding.

In summary, we present the first application of recurrent neural networks to dynamically predict need for packed red blood cell transfusion over time using electronic health record data. We report superior performance compared to the Glasgow Blatchford Score. Our approach may lead to delivery of earlier resuscitation with packed red blood cells to minimize ischemic end organ damage in patients with severe acute gastrointestinal bleeding. Future directions include external validation of the model on other cohorts of high-risk patients with gastrointestinal bleeding, along with prospective implementation and deployment in the electronic health record system for high risk patients with gastrointestinal bleeding.

## Data Availability

The MIMIC-III database repository is available at https://physionet.org/content/mimiciii/1.4/.

## Abbreviations

(RNN): Recurrent Neural Network
(LSTM): Long-Short Term Memory
(EHR): Electronic Health Record
(MIMIC-III): Medical Information Mart for Intensive Care III
(AUROC): Area Under the Receiver Operating Curve
(GBS): Glasgow-Blatchford Score

